# Optimal and Safe Pain Management Approach in Ankle and Hindfoot Fractures: Improving Practitioner Decision

**DOI:** 10.1101/2023.12.08.23299738

**Authors:** Tashfeen Ahmad, Zehra Abdul Muhammad, Yasir Mohib, Riaz Hussain, Masood Umer

**Affiliations:** Departments of Surgery and Biological & Biomedical Sciences, Aga Khan University, Karachi, Pakistan; Department of Surgery, Aga Khan University; Departments of Surgery, Aga Khan University, Karachi, Pakistan

**Keywords:** Ankle fracture, Hindfoot fracture, Trauma, Optimal pain management, Adverse events, Oral analgesics

## Abstract

**Background:** Over or sub-optimal analgesic treatment leads to undesired consequences and patient dissatisfaction. The study aims to assess the sub-optimal or optimal pain relief and safety of routinely prescribed oral analgesic(s) at discharge and 1-week post-discharge in ankle and foot fracture surgeries.

**Methods:** The ongoing prospective cohort study data on 54 ankle and hindfoot trauma fracture adult patients enrolled between June 2022 to July 2023 was analyzed. Post-surgery oral analgesics prescribed at hospital discharge and 1-week follow-up were stratified for assessing adverse events and pain (Visual Analogue Scale) at 1- and 2 weeks post-discharge. The relationship of age, gender, and comorbidity was analyzed by multiple logistic regression for adverse events and multiple linear regression for pain score.

**Results:** Median pain scores at 1- and 2-week follow-ups were 3.2 (IQR=3.0) and 2 (IQR=2.0) respectively. Combinations of tramadol, acetaminophen with naproxen or diclofenac or orphenadrine; and naproxen, pregabalin, with acetaminophen seemed toxic with sub-optimal pain control. Similar results were for celecoxib combined with pregabalin and etoricoxib combined with diclofenac or tramadol. Acetaminophen alone was safe but occasionally showed intolerance. Etoricoxib or diclofenac alone or with acetaminophen was safe and showed better pain control in this cohort. A regression model was non-significant for a relationship between covariates and pain scores or adverse events.

**Conclusion:** Current data suggests that certain oral analgesics or their combinations are harmful with sub-optimal pain control while some are safe and effective. Choosing suitable analgesics or their combinations in specific fractures might reduce patient harm with optimal pain management.

## Introduction

The ankle and hindfoot are essential anatomical parts for controlling body balance, and mobility. If fractured, there is difficulty in ambulation and fracture pain further restricts these movements. Fracture pre-and post-treatment pain control is essential to reduce patient distress, but over or sub-optimal treatment leads to undesired consequences and results in readmissions, prolonged treatment, and patient dissatisfaction. Further to this, while selecting analgesics, pain control is mainly focused, ignoring potential patient harm. Parenteral analgesics are usually switched to oral analgesics at the time of hospital discharge to control pain for ambulation and rehabilitation. To optimize pain management, it is of utmost importance to choose analgesics that timely and adequately manage pain and have negligible side effects.

There is scarce data available on optimal pain management (including pain relief as well as analgesic safety) for specific fractures in adults [1-6]. Some pain management standards like The National Institute for Health and Care Excellence (NICE) guideline, Centers for Disease Control and Prevention (CDC), and some research studies recommend acetaminophen as the first treatment choice followed by either NSAIDs or opioids depending on fracture type, site, etc. [7, 8]. In the NICE guide, pain management for ankle and foot fractures is also specified. Oral acetaminophen is recommended as an initial treatment for ankle fractures. Codeine could be added for uncontrolled pain with further addition of NSAIDs as required but with due care in elderly and frail adults [7]. Injectable morphine might be administered in uncontrolled ankle fracture pain with special attention in elderly and frail adults [7, 9]. Furthermore, certain research studies developed ankle and foot fracture pain management protocols with a reduced number of analgesic quantities while in some studies patient satisfaction with better ankle fracture pain control was assessed [10-12]. Some randomized controlled trials were performed to assess safety and pain control by certain analgesics in ankle and foot fracture surgeries [13, 14].

It is essential that practitioners have sound knowledge about appropriate analgesic selection that controls pain effectively and is safe. The study aims to assess the extent of pain relief and safety of oral analgesic(s) routinely prescribed in ankle and foot fracture surgeries at discharge and 1-week post-discharge at a tertiary care hospital. This will allow us to identify the analgesics or their combinations that either adequately control pain with better tolerance or inadequately control pain with potential toxicity. The current evidence will help maximize benefit and reduce patient harm by selecting appropriate and safe analgesics for ankle and hind foot trauma fracture pain management and thus, optimize orthopaedic surgeons analgesic prescribing practice.

## Methods

After obtaining Ethical Review Committees and Institutional approvals, the observational, prospective cohort study was initiated in June 2022. The adult patient population with ankle or hindfoot trauma fractures of any gender who arrived at a tertiary care hospital between June 2022 to July 2023 and were surgically managed were included. Those patients who were unable to consent or had undergone limb amputation were excluded. The patient’s written informed consent was obtained as per Good Clinical Practice guidelines before study enrollment. The demographics and prescribed oral analgesics at patient discharge and 1-week follow-up were recorded. Continuous variables like for age, normality of distribution was assessed for normality of distribution using the Shapiro Wilk test. For qualitative variables, frequency and percentages were used. Patients were followed at 1- and 2-week post-discharge to assess their pain control using a visual analogue scale (0=no pain, 1-3=mild, 4-6=moderate, and 7-10=severe pain scores) as well as any adverse events (reported by patients or were clinically examined/identified) likely with prescribed oral analgesics at discharge and 1-week follow-up respectively. Oral analgesics or their combinations prescribed at discharge and 1-week follow-up were stratified. Analgesics with sub-optimal pain control and toxicity or with optimal pain control and tolerance were isolated. The relationship of covariates age gender, and comorbid condition was evaluated between adverse events or pain scores assessed at 1- and 2-week follow-ups using multiple logistic regression or multiple linear regression analysis respectively. The *p*-value <0.05 was considered statistically significant with a confidence interval of 95%.

## Results

During almost one year time, 54 ankle and hindfoot fracture patients were enrolled in the study in which ankle fractures 46 (85%), hindfoot 7 (13%), and both ankle and hindfoot fractures 01 (2%). Males were 33 (61%) while females were 21 (39%). The median age was 46 (IQR=25). The most common mechanisms of injury were falls (N=24, 44%) and road traffic accidents (N=23, 43%). Median pain scores were 3.2 (IQR=3.0) and 2.0 (IQR=2.0) at 1- and 2-week follow-ups respectively.

At 1 week follow-up, 25 (46%) patients were free of any AE/SAE while 29(54%) experienced either AE (N=17) or SAE (N=12) potentially due to analgesics prescribed at the time of discharge. At 2 weeks follow-up, 38(70%) patients were free of AE/SAE while 16(30%) experienced either AE (N=12) or SAE (N=04) potentially due to 1-week prescribed analgesics. No mortality was reported at both follow-ups.

On stratification (Figures 1 and 2), the prescribed analgesics showing toxicity and with sub-optimal pain control or analgesics showing better pain control and tolerance at 1- and 2-week follow-ups were segregated. It was observed that analgesic combinations like the combination of acetaminophen, tramadol, and diclofenac (N=8) showed several SAEs (N=3) and AE (N=2) with moderate pain in half of the patients at follow-ups. Naproxen, acetaminophen, and pregabalin with or without adding thiocolchicoside or tizanidine (N=3) showed SAE (N=1) or AE (N=1) with mostly moderate pain. Adding tramadol in the analgesic combination naproxen, acetaminophen, and pregabalin (N=2) shows AE (N=1) and mild/moderate pain. Combining naproxen, acetaminophen, and tramadol (N=2) also showed AE in both patients with mild/severe pain.

**Figure 1:**
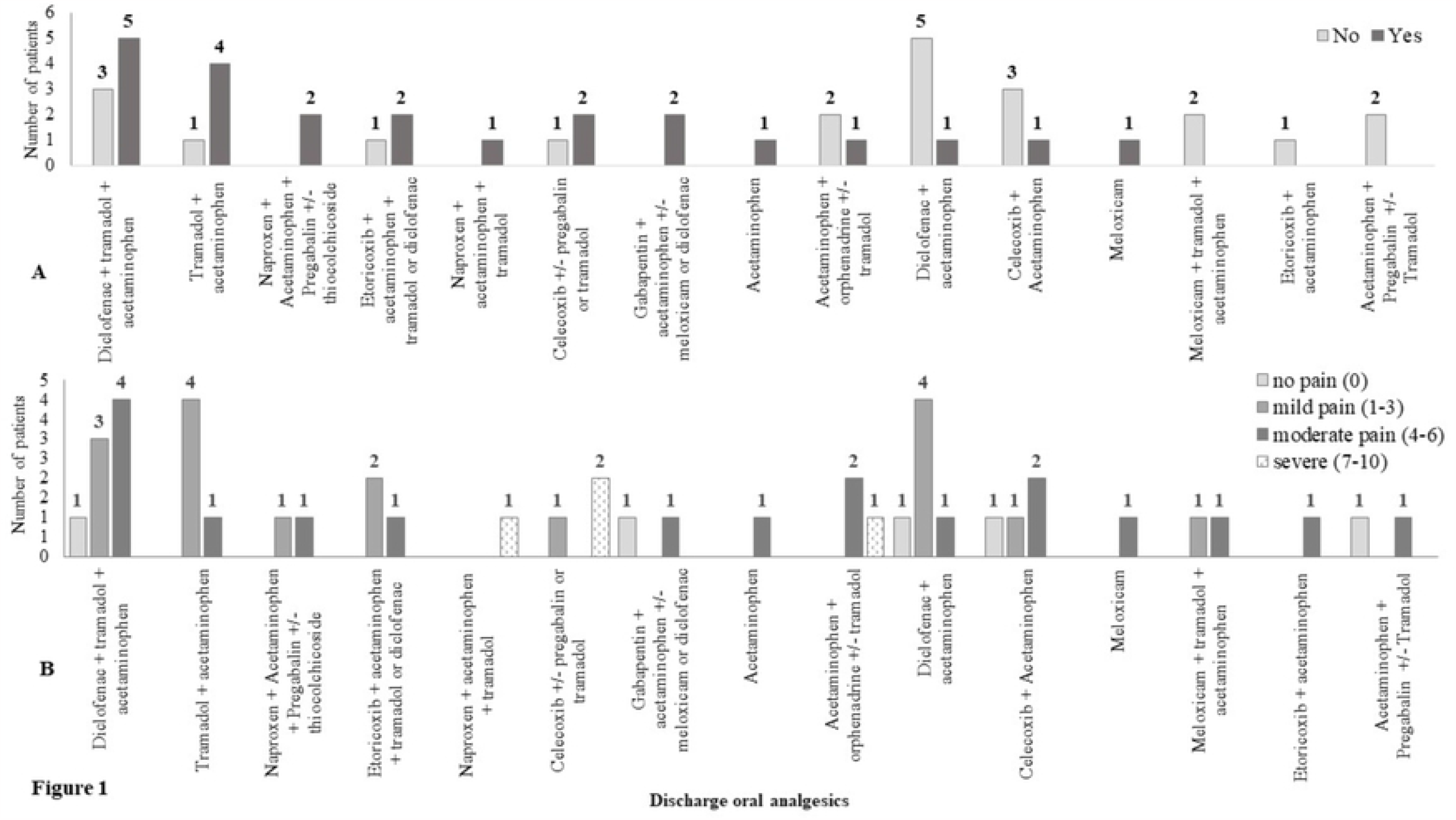
Discharge analgesics with potential SAE/AEs (A) and pain control (B).

**Figure 2:**
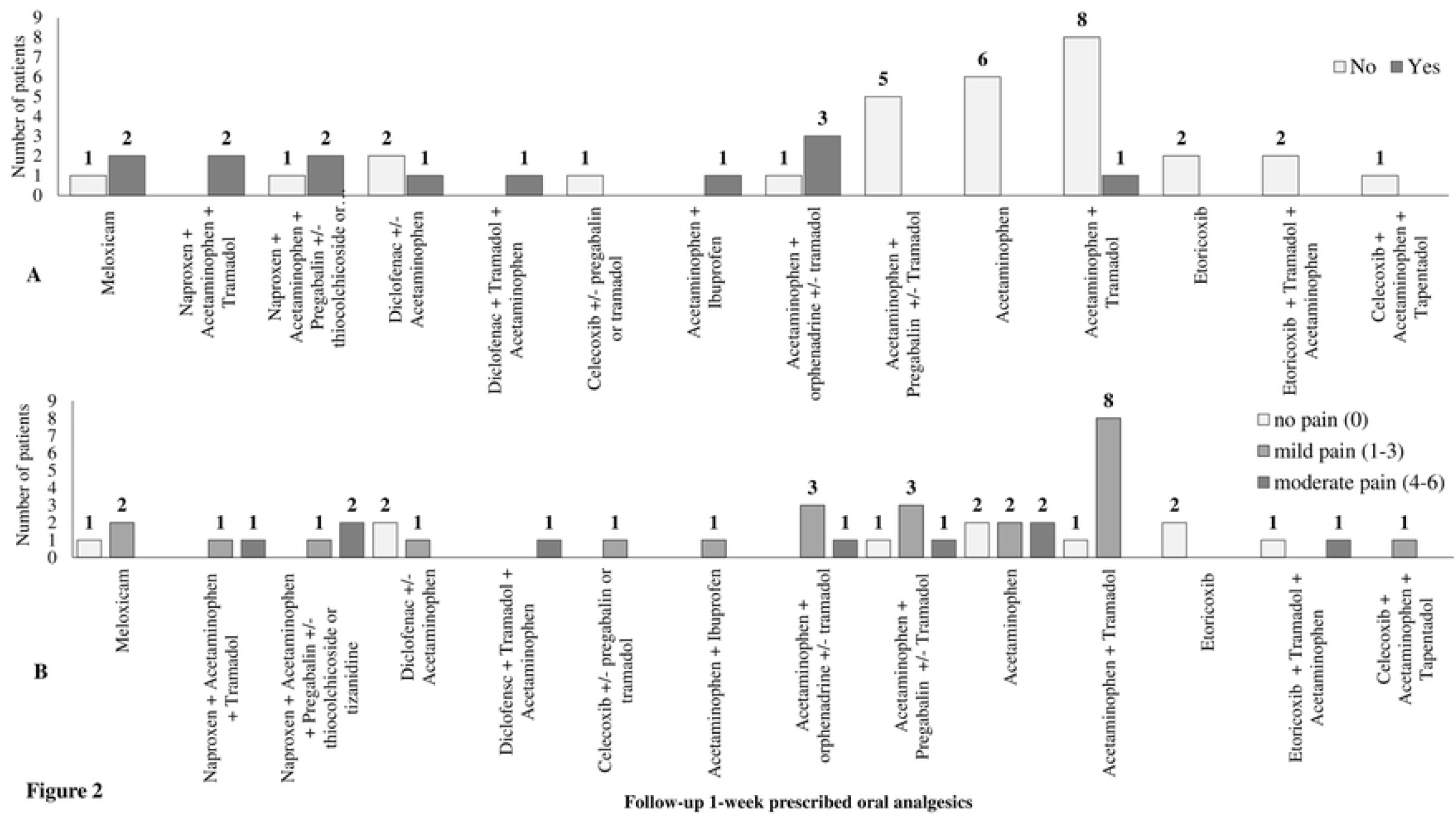
Analgesics prescribed at 1-week follow-up with potential SAE/AEs (A) and pain control (B).

Patients treated with the combination of acetaminophen and orphenadrine with or without adding tramadol (N=5) experienced SAE (N=3) and AE (N=1) with inadequate pain control in the majority at follow-ups. Combination of celecoxib, and acetaminophen (N=4) showed AE (N=2) and no to moderate pain. If celecoxib was combined with pregabalin or tramadol (N=3) showed AE (N=2) with severe pain (N=2). Meloxicam when administered alone (N=4) showed SAE (N=1) and AE (N=2) with none to moderate pain. When meloxicam was combined with tramadol and acetaminophen (N=2) showed no AE with mild/moderate pain (N=2).

Tramadol, when combined with acetaminophen (N=9) produced AEs (N=4) but pain was well controlled in these patients at follow-ups.

Some analgesics in certain analgesic combinations worked well while when combined with some other analgesics showed toxicity and inadequate pain control. Analgesic etoricoxib (N=2) had no AE/SAE and no/mild pain. Adding acetaminophen in etoricoxib (N=1) showed no AE/SAE but moderate pain. If etoricoxib was combined with tramadol or diclofenac (N=3) showed SAE (N=1) or AE (N=1) with no to moderate pain. Patients treated with diclofenac with or without acetaminophen (N=7) experienced only mild AE (N=2) with no/mild pain in the majority.

Acetaminophen (N=6) administration showed SAE (N=1) with no to moderate pain. The combination of pregabalin and acetaminophen (N=5) with or without tramadol showed no SAE/AE and pain was none or mild in the majority.

On applying multiple logistic regression, the model was non-significant for AEs assessed at 1-(X^2^(3) = 0.47, *p*=0.9) and 2 weeks (X^2^(3) = 3.9, *p*=0.26) to covariates age (*p*=0.9, 0.6), gender (*p*=0.7, 0.6), and comorbidity binary variable (*p*=0.6, 0.2) with Nagelkerke R^2^=0.01 and 0.1, respectively. The multiple linear regression model was non-significant for pain score at 1- and 2-week follow-ups and covariates age (*p*=0.1, CI= -0.67-0.13; *p*=0.9, CI= -0.03-0.03), gender (*p*=0.7, CI= -1.2-0.9; *p*=0.1, CI= -1.6-0.2), and comorbidity binary variable (*p*=0.1, CI= -0.2-2.3; *p*=0.2, CI= -0.4-1.8) with R^2^=0.05 and 0.07 respectively.

## Discussion

In the current data, it was observed that some oral analgesics prescribed to ankle or hindfoot fracture patients at discharge or at 1-week follow-up are sub-optimal in controlling pain and showed toxicity at 1- and/or 2-week follow-ups respectively. When acetaminophen, tramadol, and diclofenac were combined, several central nervous system (CNS) and cardiovascular side effects were reported like anxiety, insomnia, headache, dizziness, angina, hypotension, hypertension, etc. with sub-optimal pain control. In the literature, diclofenac risks are reported, particularly cardiovascular events [15, 16]. Additionally, some evidence shows that combining tramadol and diclofenac can cause CNS and GIT side effects but contrary to our findings, better analgesia was observed in these studies [17-19].

Another combination of naproxen, acetaminophen pregabalin with or without adding other analgesics like thiocolchicoside or tizanidine or tramadol showed sub-optimal pain control with GIT, cardiovascular (CVS), and CNS toxicity like hypotension, headache, anorexia, dysphagia, constipation, oral ulcers, diarrhea, weakness, cough etc. Similar reports are available on tramadol and naproxen toxicity and sub-optimal pain control on opioid use [20-27]. In addition to the above analgesic combinations, acetaminophen and orphenadrine including or not tramadol showed sub-optimal pain control with toxicity like hypotension, hypertension, dyspnea, sweating, dysuria, anorexia, depression, etc. There is scarce literature available on the toxicity of this combination [28].

When celecoxib and acetaminophen were combined showed better pain control in the majority and produced gastrointestinal (GIT) side effects like constipation, nausea, and vomiting. The addition of pregabalin in this combination produced CNS and GIT side effects like insomnia, constipation, diarrhea, etc. with sub-optimal pain control. Our finding is contrary to available scarce evidence of celecoxib with pregabalin being safe and effective in spinal surgery [29]. As the indication of pain management is different, the results may vary. Celecoxib toxicity is also reported for CNS, GIT, and CVS systems [30, 31].

Meloxicam when taken alone, one 30-year-old patient experienced SAE hypotension with no underlying comorbidity. Besides this, GIT-related side effects were also observed in patients like constipation and diarrhea. The pain was mostly well controlled. The side effects of meloxicam are commonly related to GIT problems, but cerebrovascular side effects are also known [32-34].

Etoricoxib when combined with acetaminophen was safe with no side effects and better pain control in ankle and hind foot fractures but when tramadol or diclofenac is added, the combination showed severe CNS toxicity like memory loss, blurred vision, fatigue, dry mouth along with GIT problems like anorexia, constipation, nausea with sub-optimal pain control. Few reports are available for this combination in which side effects of etoricoxib when combined with diclofenac are mentioned like GIT and CNS problems including seizures, hypertension, etc. [35-38] Analgesic pregabalin alone or with acetaminophen and tramadol showed no AE with none to moderate pain. Rare reports are available for this combination which showed better pain control with AEs like somnolence, dizziness, and blurred vision [39].

In the literature, acetaminophen is reported to be a safe analgesic [40, 41]. In the present research, mostly there was no harm observed after acetaminophen administration except for one 24-year-old pregnant lady with no underlying comorbidity who experienced hypotension. It was also observed that pain control was sub-optimal with acetaminophen alone. The literature reports similar systemic events including hypotension after taking acetaminophen [42-48].

In our ankle and hindfoot fracture population, analgesic diclofenac with or without acetaminophen optimally controlled pain as well as was safe in ankle and hindfoot fractures. When diclofenac was combined with tramadol showed SAE/AEs but when prescribed alone or in combination with acetaminophen, it seemed safe and had better pain control.

## Conclusion

Thus, in the current ankle and hindfoot fractures data on oral analgesics management, it was observed that certain analgesic combinations were sub-optimal in controlling pain and were toxic like combinations of tramadol with diclofenac/naproxen; naproxen, pregabalin and acetaminophen; etoricoxib with tramadol/diclofenac; orphenadrine with acetaminophen with or without tramadol; and celecoxib with acetaminophen with or without tramadol/pregabalin. Changing some sets of analgesic combinations were safe and had better pain control like etoricoxib or diclofenac with or without combining with acetaminophen showed better tolerance and pain control in ankle and hindfoot fracture patients. The current stratification of harmful and safe analgesic combinations might assist in optimal pain management of specific fractures by selecting safe analgesics that have better pain control.

## Limitation

This is a single-center study and thus, the generalizability of findings was not achieved. The analgesics to AE relationship was not determined thoroughly by root cause analysis.

## Disclosure

The authors report no proprietary or commercial interest in any product mentioned or concept discussed in this article.

## Funding

The study was generously supported by the Orthopaedic Trauma Association (proposal ID #6704, https://ota.org/). The award was received by TA, ZM, and YM.

## Conflict of interest

The Authors declare that there are no conflicts of interest regarding the publication of this article.

## Data availability

On reasonable request, data will be provided.

## Notes

### Competing Interest Statement

The authors have declared no competing interest.

### Funding Statement

Yes

### Author Declarations

National Bioethics Committee and Aga Khan University Ethical Review Committee approvals were obtained prior to study start-up.

